# Socioeconomic inequalities in cannabis use among European adolescents: the role of access and risk perception

**DOI:** 10.64898/2026.09.01.26361911

**Authors:** Julian Perelman, Márcia Cardoso, Joana Alves, Claudia Palladino

## Abstract

**Introduction:** There is evidence that regular and high-risk-consumption is more prevalent among adolescents with a lower socioeconomic family status (SES). Yet, the causes for the SES gap remain unclear. We postulate that SES inequalities in regular and high-risk cannabis use among adolescents are mediated by differences in risk perception and access.

**Methods:** This cross-sectional study used data from a survey to students aged 15-17 years from 35 European countries in 2019 (European School Survey Project on Alcohol and Drugs). We modelled current regular use (≥10 times in the past month) and past-year high-risk/problematic use as a function of maternal and paternal education, adjusting for age, sex, and country, then further adjusting for moderating variables: perceived cannabis availability and perceived risk of harm.

**Results:** Lower paternal education was related to both higher regular and high-risk use (OR: 1.55 (1.20-2.0) and OR: 1.77 (1.49-2.11), respectively. A similar pattern was observed with maternal educational level. Including mediating factors, perceived availability increased the likelihood of higher use among adolescents with lower SES, related to their lower perceived availability. In contrast, adjusting for risk perception, SES factors lost significance for regular use and reduced OR by 40 to 50% for high-risk use.

**Conclusion:** Results indicate that SES inequalities in perceived risk explain the SES gap in regular and high-risk cannabis use among the worse-off. Attention should be paid to policies that may foster the perception that cannabis is risky, which may aggravate dangerous consumption among the worse-off, contributing to larger SES inequalities.

## INTRODUCTION

Adolescence is a crucial period of cognitive, psychological and social development. These changes lead to an interest in new sensations and experiences, making it a high-risk period for drug initiation (1). The adolescent brain’s exposure to the lasting negative effects of early drug use leads to a more rapid development of addiction and potential problems in adulthood (2). As an illicit drug, the case of cannabis is of primary relevance for global public health (3,4). Indeed, cannabis is the most used illicit drug among the general population and among young people worldwide. In 2021, the annual prevalence of cannabis use was 5.3% among the population aged 15-16 years (13.5 million persons), as compared with 4.3% among the population aged 15-64 years (3). Cannabis is also the most commonly used illicit substance in the European Union, with a prevalence of last year use of 8% among European adults (22.6 million aged 15-64), with peaks among young people, i.e., 15% among those aged 15-34, 18% among the 15-24, and 13% among adolescents aged 15-16 (5,6).

The non-medical use of cannabis is prohibited by international control regulations due to the health risks associated with its use (7), mainly due to the presence of the major cannabinoids delta-9-tetrahydrocannabinol and cannabidiol. The main adverse outcomes are mental illnesses, cannabis use disorder, acute cognitive and psychomotor control impairment (8). Furthermore, cannabis use increases the risk of interpersonal violence and of traffic accidents (9). The inhalation and emission of fine particles and carcinogens from smoking cannabis, the most common method of using this substance, causes damage similar to that caused by tobacco (10). Recently, new cannabis policies have being developed in Europe to address not only the control of illicit cannabis, but also the regulation of cannabis and cannabinoids for therapeutic and other uses. The health and social consequences of all these issues cannot be overlooked as a public health concern (11).

In addition to the inherent characteristics of adolescent development that may facilitate the initiation of drug use, other factors play a role at these ages. These include peer influence, parental drug use, patterns of family dysfunction, abuse, low self-esteem, and poor academic performance, which have been identified as associated with increased drug use among adolescents (12,13). The socioeconomic status (SES) has also been identified as an important determinant of youth drug use (12–14), although results are contradictory. A review of 2007 found no clear pattern of association between parental education, an indicator of SES, and cannabis use (15), while more recent evidence indicates an association between low SES and cannabis use (16,17). Notably, trends in cannabis use among Finnish adolescents with most socioeconomic challenges increased significantly between 2000 and 2015 (18). According to some authors, cannabis use is more prevalent among adolescents from higher SES, although heavy use occurs primarily in disadvantaged contexts (13,17,19,20). In addition to ambiguous evidence, the mechanisms underlying the relationship between SES and cannabis use remain poorly understood.

Notably, attitudes towards cannabis use may influence adolescent use, including perceived availability and harmfulness. The concept of perceived availability is inherently subjective, and the perception of risk is sufficiently complex to include the perception of the likelihood of direct and indirect, short-term and long-term consequences (21). Perceived harmfulness is a protective factor against its use (22). Traditionally, cannabis access is seen as low, and risk high due to its illegal status. However, with increasing legalisation, easy access and low risk perception can lead to unpredictable behaviour. Evidence shows a decline in risk perception after cannabis policy changes in the US (22,23), along with increased use among both adolescents and young adults and a concurrent decrease in perceived risk between 2008–2019 after recreational legalization in this country (24). Perceived risk and availability may also have a positive additive interaction for cannabis use, being the combined effect of perceiving cannabis as low-risk and available on past-year use higher than the sum of each factor individually (25).

In this paper, we postulate that socioeconomic inequalities in regular and high-risk cannabis use among adolescents are mediated by differences in risk perception and access. By doing so, we search for a better understanding of the socioeconomic differences in cannabis use among adolescents, which may help support decision making in the field, namely whether policies reducing easy access or informing about risk might help mitigate inequalities against the worse-off.

## PARTICIPANTS AND METHODS

### Study design and population

A cross-sectional study was conducted using data from the European School Survey Project on Alcohol and Drugs (ESPAD) 2019 (5). The target population was defined as students who reached the age of 16 years in the calendar year of the survey and who were present in the classroom on the day of the survey. Participants were selected using a multistage stratified random sampling procedure. The ESPAD questionnaire was administered anonymously to 102,484 students aged 15-17 years from 35 European countries (26). All samples were nationally representative, except those from Cyprus, Kosovo, Georgia, and Germany. After removing inconsistent or insufficiently completed questionnaires, a total of 101,694 observations remained for analyses. The final sample sizes for each of the participating countries are shown in Supplementary Table 1. This study did not require ethical approval or informed consent, as it is a secondary analysis of data that was previously collected on an anonymous basis.

### Measures

#### Outcomes

The dependent variables were used to characterise adolescents’ cannabis use. These included current regular use (defined as consumption of ten or more times in the past month) (27), and past-year high-risk/problematic use. For the latter, participants were considered at high risk of cannabis-related problems if they had used cannabis in the past 12 months and scored 2 or higher on the Cannabis Abuse Screening Test (CAST) (28,29). The CAST index was developed by researchers in 2007 and subsequently adopted by the European Monitoring Centre for Drugs and Drug Addiction (EMCDDA) to assess the extent of cannabis-related problems (30,31). The CAST score is based on six items relating to the previous year, each with a five-point response scale ranging from ‘never’ to ‘very often’ (26). All items were re-categorised into ‘0’ and ‘1’. The CAST score was then calculated as the sum of the six CAST items, ranging from 0 to 6. A binary recoding was then used, with a cut-off of 2 or more points, to distinguish students at high risk of cannabis-related problems from those at low risk of such problems. For a detailed description of all measures included, as well as the questions as reported in the ESPAD questionnaire and the codes used, see Supplementary Table 2.

#### Explanatory variables

The SES indicators used as explanatory variables were represented by the educational levels of the mother and father, with each parent’s highest level of education considered as a separate measure. This variable is commonly used to assess SES among adolescents (20). Educational attainment was divided into five categories, ranging from “completed primary school or less” to “completed college or university”.

#### Mediators

The mediating variables were the perceived availability of cannabis, assessed in terms of the ease of obtaining cannabis (divided into four categories, from “impossible or very difficult” to “very easy”), and the perceived risk of harm associated with experimental, occasional and regular cannabis use. These were divided into four categories (“no risk”, “slight risk”, “moderate risk”, “great risk”).

#### Covariates

Sex and country were included because of their association with the outcome variables. The previous ESPAD survey (2015) showed that male adolescents generally have a higher prevalence of cannabis use (32). In addition, a number of heterogeneous political and cultural factors may influence consumption patterns across countries, and use varied significantly across the ESPAD countries (32). The age range of 15-17 years was an inclusion criterion for respondents, and thus age was not included in the analysis.

### Model

In accordance with the model proposed in this study, the SES (explanatory variable, X) exerts an influence on cannabis use (dependent variable, Y), and this relationship is itself subject to the influence of risk perception and access perception (mediating variables, M). As previously stated, sex and country were considered covariates.

### Statistical analysis

The prevalence and frequency of cannabis use and a descriptive overview of the other variables are presented using descriptive statistics. In the bivariate analysis, the Pearson chi-square tests were used to compare categorical variables. The association of both SES (explanatory) and availability and risk perception (potential mediators) with cannabis use (outcome) was assessed separately by analysing the prevalence of cannabis use within each SES and mediating category by Pearson chi-square tests. The results of the Pearson chi-square tests were confirmed by univariate logistic regression. The association between SES and availability and risk perceptions was also assessed, analysing the distribution of each perception level within each SES category. Only when availability and risk perception variables are associated with both SES and cannabis use can they play a role as mediators.

The mediation analysis consisted of three sets of regressions. In a first regression (model 1) - the association of each SES (as indicated by education) with each outcome (regular use, and high-risk use) was examined separately using binary logistic regressions. Prevalence differences between SES groups were expressed in odds ratios (ORs) with 95% confidence intervals (CI). The highest level of education and of family well off were used as references.

In a second regression (model 2) - the association of SES (as indicated by education) with each of the availability perception as mediator was assessed using generalised linear models.

Third regression – all potential mediators were added to model 1, resulting in logistic regressions with the three types of variables simultaneously - explanatory, mediator, and outcome. All models were adjusted for country and sex as confounders.

Full mediation occurs when the explanatory variable no longer has a significant link after the mediator is introduced into the equation. Odds ratios (ORs) of the various associations were compared. The observed reduction in the strength of the association between SES and cannabis use can be attributed to the influence of availability and risk perception. The percentage reduction of the effect was calculated in accordance with the methodologies previously established in the literature (33–35): (OR _model 1_ – OR _adjusted model_)/(OR _model 1_ – 1) × 100%.

The percentage reduction in OR was not analysed for associations that showed non-significant coefficients or where the OR changed direction, i.e., with intervals between OR_model 1_ – OR_adjusted model_ containing the value 1. P-value *<* 0.05 (2-sided) was considered statistically significant; SPSS (v.28; Chicago, IL) and R v.2023.12 were used.

## RESULTS

### Study population characteristics

A total of 101,694 participants were included, with country sample sizes ranging from 417 in Monaco (0.4% of the sample) to 5,942 in Greece (5.8%) (Supplementary Table 1). Most participants were 16 years old (96.6%), and a majority were female (51.4%) (Table 1). A fifth of fathers (20.4%) did not complete secondary school, for 15.8% of the mothers. A high proportion of adolescents (39.2%) reported cannabis to be easily available (‘fairly easy’ or ‘very easy’). More than half of the participants perceived ‘no risk’ or ‘slight risk’ to ‘try cannabis once or twice’ (53.2%). This proportion decreased for more intense use, i.e. for ‘occasional use’ (33.0%) and ‘regular use’ (13.7%).

**Table 1.** Sample description.

|  | **N** | **%** |
| --- | --- | --- |
| **Sex** |  |  |
| Male | 49 394 | 48.6 |
| Female | 52 300 | 51.4 |
| **Father education** |  |  |
| Completed primary school or less | 6 580 | 8.0 |
| Some secondary school | 10 235 | 12.4 |
| Completed secondary school | 27 764 | 33.7 |
| Some college or university | 11 077 | 13.4 |
| Completed college or university | 26 827 | 32.5 |
| **Mother education** |  |  |
| Completed primary school or less | 5 591 | 6.5 |
| Some secondary school | 8 064 | 9.3 |
| Completed secondary school | 26 669 | 30.8 |
| Some college or university | 12 498 | 14.4 |
| Completed college or university | 33 758 | 39.0 |
| **Perceived availability** |  |  |
| Impossible or very difficult | 37 255 | 45.9 |
| Fairly difficult | 12 082 | 14.9 |
| Fairly easy | 19 653 | 24.2 |
| Very easy | 12 105 | 14.9 |
| **Perceived risk (try once or twice)** |  |  |
| No risk | 20 300 | 22.9 |
| Slight risk | 26 887 | 30.3 |
| Moderate risk | 19 060 | 21.5 |
| Great risk | 22 472 | 25.3 |
| **Perceived risk (occasional use)** |  |  |
| No risk | 9 988 | 11.3 |
| Slight risk | 19 222 | 21.7 |
| Moderate risk | 30 012 | 33.8 |
| Great risk | 29 513 | 33.3 |
| **Perceived risk (regular use)** |  |  |
| No risk | 5 638 | 6.3 |
| Slight risk | 6 536 | 7.3 |
| Moderate risk | 16 672 | 18.7 |
| Great risk | 60 130 | 67.6 |
| **Current regular use (among users in last month)** |  |  |
| Not regular | 5 271 | 80.0 |
| Regular | 1 315 | 20.0 |
| **High-risk /problematic use (among users in last year)** |  |  |
| No high-risk (CAST<2) | 7 615 | 66.2 |
| High risk (CAST 2+) | 3 888 | 33.8 |

Among those with current use (n=6,586; 6.5% of the sample), the prevalence of regular use was 20.0%. Regarding the risky levels of cannabis use, among the overall ESPAD sample of both past-year users (n = 11,503) and non-users (n = 89,107) and available information on the CAST items, 4.0% (n=3,888) of students were classified as high-risk/problematic users. However, this prevalence increased to 33.8% among those who had consumed cannabis in the past 12 months.

### Bivariate analyses

Lower father’ education was related to both higher regular and high-risk use, with prevalence of 24.2% [OR: 1.55 (1.20-2.0)] and 41.9% [OR: 1.77 (1.49-2.11)] (Supplementary Table 3). A similar pattern was observed in the association between mother’s educational level and cannabis use. We observed an increased prevalence and likelihood of cannabis use among those who perceived it as very easy to obtain [27.3%, OR: 1.53 (1.13-2.07); 44.0%, OR: 2.24 (1.84-2.73) for regular and high-risk use, respectively]. The prevalence and likelihood of cannabis use were generally higher among those who did not perceive any risk of harm than among those who perceived high risk, peaking at 49.7% [OR: 2.65 (2.35-2.99)] of those who perceived no risk of harm from regular use engaging in high-risk use.

Overall, the proportion of those who perceived cannabis to be easily available (‘fairly easy’ or ‘very easy’) increased with higher levels of parental education (42% for the group whose mothers or fathers had ‘completed college or university’) (Supplementary Table 4). The proportion of respondents who perceived no or slight risk of trying cannabis once or twice was highest in the group whose parents had the highest educational level (55.8%). Notably, this pattern reversed for those who perceived no or slight risk of trying cannabis regularly, being those whose parents had the lowest educational level more represented (16-17%).

### Multivariate analyses

After adjusting for sex and country (model 1), the odds of regular and high-risk use steadily increased with decreasing levels of education (Table 2). Respondents whose parents completed primary school or less were 62-70% more likely to engage in regular use and 76-90% more likely to engage in high-risk use compared to respondents whose parents completed college or university.

**Table 2.** Association between current regular/high-risk cannabis use and socioeconomic variables, adjusted for sex and country (Model 1)

|  | Regular use |  |  | High-risk use |  |  |
| --- | --- | --- | --- | --- | --- | --- |
|  | OR | 95% CI | *p-value* | OR | 95% CI | *p-value* |
| **Explanatory** |  |  |  |  |  |  |
| **Father education** |  |  |  |  |  |  |
| Total |  |  |  |  |  |  |
| Completed college or university | 1 |  |  | 1 |  |  |
| Completed primary school or less | 1.62 | (1.24-2.11) | <0.001 | 1.90 | (1.59-2.28) | <0.001 |
| Some secondary school | 1.34 | (1.07-1.69) | 0.012 | 1.51 | (1.31-1.75) | <0.001 |
| Completed secondary school | 1.13 | (0.94-1.35) | 0.201 | 1.27 | (1.14-1.43) | <0.001 |
| Some college or university | 0.99 | (0.79-1.24) | 0.926 | 1.22 | (1.06-1.41) | 0.005 |
| **Mother education** |  |  |  |  |  |  |
| Total |  |  |  |  |  |  |
| Completed college or university | 1 |  |  | 1 |  |  |
| Completed primary school or less | 1.70 | (1.26-2.29) | 0.001 | 1.76 | (1.44-2.16) | <0.001 |
| Some secondary school | 1.41 | (1.11-1.8) | 0.006 | 1.59 | (1.36-1.86) | <0.001 |
| Completed secondary school | 1.22 | (1.03-1.45) | 0.025 | 1.25 | (1.12-1.39) | <0.001 |
| Some college or university | 1.16 | (0.94-1.42) | 0.167 | 1.13 | (1.00-1.29) | 0.058 |

When the total effect model was adjusted for the perceived availability, the odds of regular and high-risk/problematic use for the disadvantaged adolescents whose parents had low education increased (Tables 3 and 4; Supplementary Figure 1). This is explained by the lower perceived availability among the worse-off; if the worse-off had the perceived availability of the better-off, their consumption would be higher, and socioeconomic inequalities would be greater.

**Table 3.** Association between current regular cannabis use and socioeconomic variables, adjusted for sex, country, and mediating variables.

|  | **Perceived availability-adjusted OR**<br>**(Model 2)** | **% variation** | **Fully adjusted OR (Model 3)** | **% variation** |
| --- | --- | --- | --- | --- |
| **Father's education** |  |  |  |  |
| Completed college or university | 1 |  | 1 |  |
| Completed primary school or less | 1.634 (1.235-2.160) | +2.69 | 1.201 (0.848-1.699) | -24.14 |
| Some secondary school | 1.442 (1.135-1.832) | +28.68 | 1.198 (0.897-1.599) | -10.57 |
| Completed secondary school | 1.200 (0.992-1.451) | +56.96 | 1.167 (.931-1.463) | -17.47 |
| Some college or university | 1.007 (0.796-1.273) | NA | .967 (0.742-1.262) | NA |
| **Mother's education** |  |  |  |  |
| Completed college or university | 1 |  | 1 |  |
| Completed primary school or less | 1.738 (1.268-2.382) | +5.51 | 1.583 (1.058-2.367) | -7.99 |
| Some secondary school | 1.474 (1.146-1.895) | +15.70 | 1.371 (1.003-1.873) | -3.87 |
| Completed secondary school | 1.250 (1.045-1.496) | +14.08 | 1.150 (0.921-1.436) | -9.51 |
| Some college or university | 1.152 (0.930-1.426) | NA | 1.178 (0.918-1.513) | NA |

NA: not available

**Table 4.**
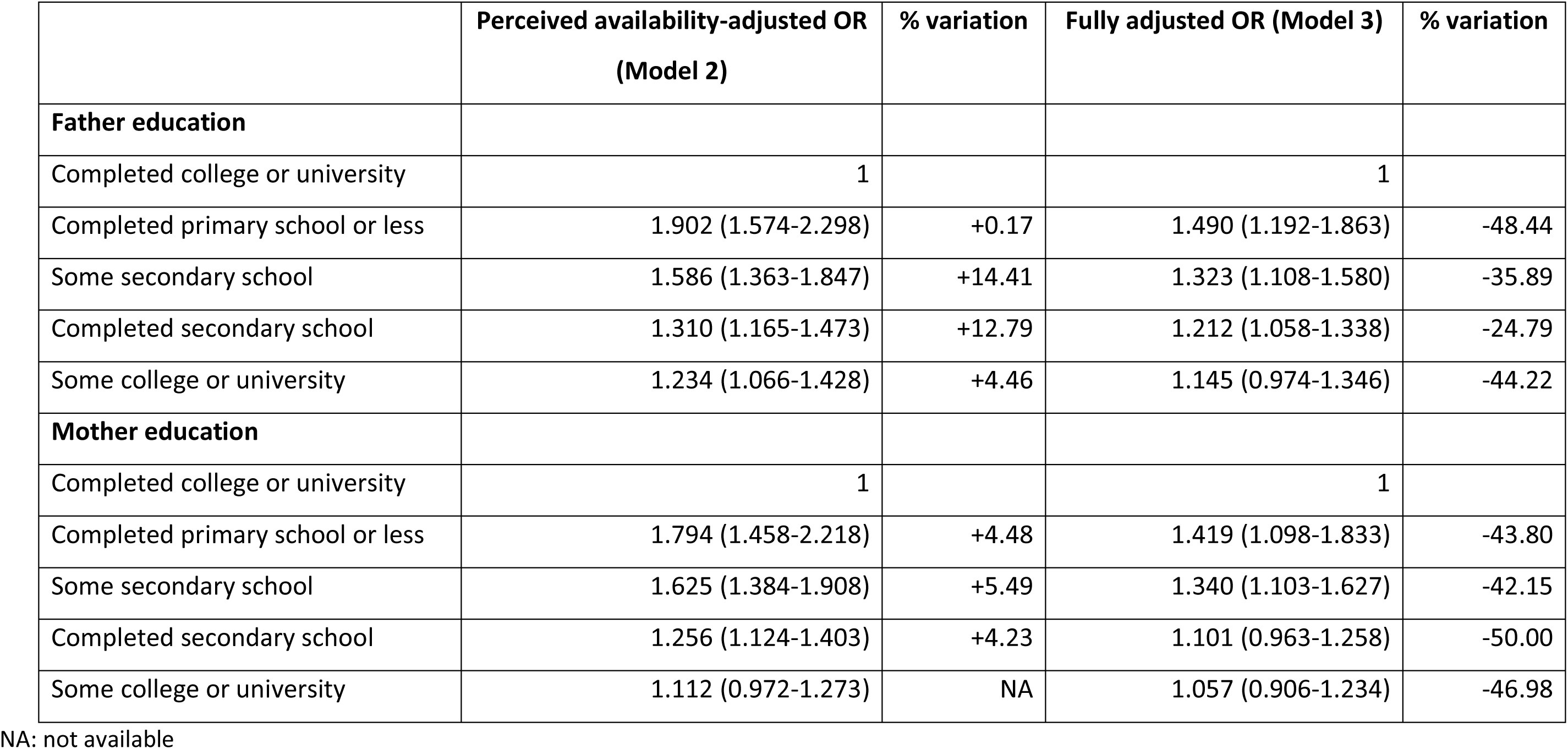
Association between past year high-risk cannabis use and socioeconomic variables, adjusted for sex, country, and mediating variables.

|  | **Perceived availability-adjusted OR**<br>**(Model 2)** | **% variation** | **Fully adjusted OR (Model 3)** | **% variation** |
| --- | --- | --- | --- | --- |
| **Father education** |  |  |  |  |
| Completed college or university | 1 |  | 1 |  |
| Completed primary school or less | 1.902 (1.574-2.298) | +0.17 | 1.490 (1.192-1.863) | -48.44 |
| Some secondary school | 1.586 (1.363-1.847) | +14.41 | 1.323 (1.108-1.580) | -35.89 |
| Completed secondary school | 1.310 (1.165-1.473) | +12.79 | 1.212 (1.058-1.338) | -24.79 |
| Some college or university | 1.234 (1.066-1.428) | +4.46 | 1.145 (0.974-1.346) | -44.22 |
| **Mother education** |  |  |  |  |
| Completed college or university | 1 |  | 1 |  |
| Completed primary school or less | 1.794 (1.458-2.218) | +4.48 | 1.419 (1.098-1.833) | -43.80 |
| Some secondary school | 1.625 (1.384-1.908) | +5.49 | 1.340 (1.103-1.627) | -42.15 |
| Completed secondary school | 1.256 (1.124-1.403) | +4.23 | 1.101 (0.963-1.258) | -50.00 |
| Some college or university | 1.112 (0.972-1.273) | NA | 1.057 (0.906-1.234) | -46.98 |

NA: not available

In the full model, additionally accounting for the perceived risk, all education variables lost significance in the case of regular use. This means that the education-related differences are fully explained by perceived risk. In the case of high-risk consumption, mediation effects were strong (ORs decrease between 24 and 50%), but the greater consumption among adolescents from low-educated parents remain significant.

## DISCUSSION

This study aimed to investigate the potential mediating role of differences in risk perception and access in the relationship between socioeconomic status and self-reported cannabis regular and high-risk use outcomes among European adolescents.

Utilising data from the most comprehensive school-based cohort on alcohol and drug use, this research addresses a critical gap, as few studies have examined the causal pathway from socioeconomic factors and cannabis use among adolescents.

The findings confirm that SES is linked cannabis use, with greater consumption among adolescents from low-educated parents. Results indicate that perceived access is greater among adolescents from more educated families; this partly attenuates the inequalities against the worse-off. In contrast, risk perception fully explains socioeconomic inequalities in regular cannabis use, and largely in high-risk use.

The study confirms that adolescent with lower SE background have greater odds of having engaged in more intense cannabis use, being almost two-fold the odds of high-risk use. This is consistent with several prospective studies highlighting association between low SES and higher rates of cannabis use and cannabis use disorder among adolescents (36,37). Some authors suggest that while cannabis use is more prevalent among adolescents from higher SES, heavy use occurs primarily in disadvantaged contexts (13,17,19,20).

Notably, limited research has explored the mediation role of individual perceptions. Studies suggest that risk perception is associated to cannabis use. A cross-sectional study of Hispanic college students in the U.S. found that higher perceived risk was linked to lower usage rates, while subjective social status was inversely related to lifetime cannabis use and positively associated with risk perception (38). Furthermore, a Finnish study of adolescents highlighted that cannabis use trends from 2000 to 2015 varied with socioeconomic adversity. Although it did not directly assess risk perception, the findings imply that adolescents facing these challenges may exhibit different usage patterns, possibly shaped by their risk and harm (18). Analysis of the ESPAD cohort confirmed that individuals with higher educational background perceive greater risks in more intense use of cannabis, contributing to reduced high-risk use in this group of adolescents. Consequently, risk perception fully mediates (i.e., explains) the SES and regular use relationship, and partially the relationship with high-risk use.

The strengths of the study include a Europe-wide representative sample with a high participation rate. Moreover, by specifically surveying 15–16-year-old adolescents, ESPAD captures critical data during a formative period for substance use, offering insight that can inform early intervention programs. The study’s limitations include reliance on parental educational as the solely measure of SES, without considering, e.g., parents’ occupational or income categories, which may reveal a more complete picture of the socioeconomic background. Note that measuring adolescent’s SES is challenging since their final education and income levels have not yet been attained. As a result, parental SES is frequently used as a proxy for adolescent SES (20).

Additionally, factors such as family structure and parenting styles, available in the ESPAD questionnaire and known to influence drug use outcomes, including paternal presence or maternal authoritative style (39), where not analysed. These parental influences should be understood within the broader social context shaping adolescent behavior. Expanding the analysis to include factors such as subjective social status, individuals’ self-perceived social standing, peer influence and school connectedness is essential, as previous studies have shown that individuals’ self-perceived social standing and peer influence plays a significant role in cannabis use in youth and that risk behaviors are associated with school absenteeism (40–42). These additions would offer a more comprehensive understanding of drug use determinants.

In conclusion, this study suggests that individuals’ perceptions of cannabis-related risks partially or fully explain the relationship between socioeconomic status and intense cannabis use. Consequently, attention should be paid to policies that might reduce the risk perception, because this reduced risk perception may be better among adolescents from worse-off families, aggravating SE inequalities. Note that, although scarce, the literature indicates that risk perception does not seem to be affected by the introduction of cannabis as medical prescription (43), nor by the legalization of cannabis for recreational use (44,45).

## Data Availability

All data produced in the present study are available upon reasonable request to the ESPAD group.

## Acknowledgments

The authors alone are responsible for the views expressed in this article and they do not necessarily represent the views, decisions or policies of institutions with which they are affiliated. The authors would like to acknowledge the members of the ESPAD group who collected the national data (http://www.espad.org/report/acknowledgements) and the funding bodies who supported the international coordination of ESPAD: the Italian National Research Council and the European Union Drugs Agency (EUDA). Special thanks are due to the schoolchildren, teachers and national funding bodies who made this project possible. ESPAD data use consent N. 2022_03_005.

## Funding

This study has received no specific funding.

## Authors’ Contributions

MC conducted the preliminary analyses. CP performed the complete analytical workflow and coordinated the study. MC and CP drafted the manuscript. JP conceptualized and coordinated the study. JA facilitated data acquisition by liaising with the survey coordinators. All authors contributed to the study design, interpretation of the results, critically revised the manuscript for important intellectual content, and approved the final version for publication.

## Notes

### Competing Interest Statement

The authors have declared no competing interest.

### Author Declarations

European School Survey Project on Alcohol and Drugs (ESPAD) data use consent N. 2022_03_005. This study did not require ethical approval or informed consent, as it is a secondary analysis of data that was previously collected on an anonymous basis.

