## Supplementary Tables and Figure for "Socioeconomic inequalities in cannabis use among European adolescents: the role of access and risk perception": SupplTabFig_as.submitted_MEDRXIV_Palladino.pdf

**Supplemental Table 1. Sample sizes by country**

| Countries | N | % |
|---|---|---|
| Austria | 4 334 | 4.3 |
| Bulgaria | 2 830 | 2.8 |
| Croatia | 2 758 | 2.7 |
| Cyprus | 1 204 | 1.2 |
| Czechia | 2 770 | 2.7 |
| Denmark | 2 488 | 2.4 |
| Estonia | 2 504 | 2.5 |
| Faroes | 510 | 0.5 |
| Finland | 4 586 | 4.5 |
| France | 2 566 | 2.5 |
| Georgia | 2 942 | 2.9 |
| Greece | 5 942 | 5.8 |
| Hungary | 2 406 | 2.4 |
| Iceland | 2 502 | 2.5 |
| Ireland | 1 929 | 1.9 |
| Italy | 2 519 | 2.5 |
| Latvia | 2 728 | 2.7 |
| Lithuania | 2 386 | 2.3 |
| Malta | 3 029 | 3.0 |
| Monaco | 417 | 0.4 |
| Montenegro | 5 687 | 5.6 |
| Netherlands | 1 284 | 1.3 |
| Norway | 4 232 | 4.2 |
| Poland | 5 019 | 4.9 |
| Portugal | 4 359 | 4.3 |
| Romania | 3 737 | 3.7 |
| Serbia | 3 492 | 3.4 |
| Slovakia | 2 244 | 2.2 |
| Slovenia | 3 398 | 3.3 |
| Spain | 3 546 | 3.5 |
| Sweden | 2 532 | 2.5 |
| Ukraine | 2 713 | 2.7 |
| North Macedonia | 2 914 | 2.9 |
| Kosovo | 1 737 | 1.7 |
| Germany (Bavaria) | 1 450 | 1.4 |
| **Total** | **101 694** | **100.0** |

**Supplemental Table 2. Variables construction and definition.**

| VARIABLES | QUESTIONS | CODES |
|---|---|---|
| **Country** | | Numeric Country Code |
| **Sex** | | Male (1) vs.<br>Female (2) |
| **Father's education** | - What is the highest level of schooling your father completed? (Question C49) | Completed primary school or less (1)<br>Some secondary school (2)<br>Completed secondary school (3)<br>Some college or university (4)<br>Completed college or university (5) |
| **Mother's education** | - What is the highest level of schooling your mother completed (Question C50) | Completed primary school or less (1)<br>Some secondary school (2)<br>Completed secondary school (3)<br>Some college or university (4)<br>Completed college or university (5) |
| **Current regular cannabis use** (among those who used during the last 30 days) | - On how many occasions (if any) have you used cannabis during the last 30 days? (Question C25c) | Not-regular use – less than 10 times (0) vs.<br>regular use – 10 times or more (1) |
| **Past-year problematic cannabis use/high-risk**<br>(among those who used cannabis in the last 12 months) | Participants considered as problematic users were those who consumed cannabis in the last 12 months and scored 2 or higher in the Cannabis Abuse Screening Test (CAST). The six items of the CAST refer to the past 12 months as follow:<br>1- Have you smoked cannabis before midday? (Question C27a)<br>2- Have you smoked cannabis when you were alone? (Question C27b)<br>3- Have you had memory problems when you smoke cannabis? (Question C27c)<br>4- Have friends or members of your family told you that you ought to reduce your cannabis use? (Question C27d) | Not high-risk use (0) vs.<br>high-risk use (1)<br><br>Codes for questions 1 and 2:<br>Never or rarely (0) vs. from time to time, fairly often, or very often (1)<br><br>Codes for questions 3-6:<br>Never (0) vs. rarely. from time to time, fairly often, or very often (1) |

| | | |
|---|---|---|
| | 5- Have you tried to reduce or stop your cannabis use without succeeding? (Question C27e)<br>6- Have you had problems because of your use of cannabis (arguments. fights. accidents. bad results at school. etc.)? (Question C27f) | |
| **Perceived availability** | - How difficult do you think it would be for you to get cannabis if you wanted? (Question C24) | Impossible or very difficult (1)<br>fairly difficult (2)<br>fairly easy (3)<br>very easy (4) |
| **Perceived risk of harm**<br>**(**try once or twice) | - How much do you think people risk harming themselves (physically or in other ways). if they try cannabis once or twice? (Question C36a) | No risk (1)<br>slight risk (2)<br>moderate risk (3)<br>great risk (4) |
| **Perceived risk of harm**<br>**(**occasional use) | - How much do you think people risk harming themselves (physically or in other ways). if they smoke cannabis occasionally? (Question C36b) | No risk (1)<br>slight risk (2)<br>moderate risk (3)<br>great risk (4) |
| **Perceived risk of harm**<br>**(**regular use) | - How much do you think people risk harming themselves (physically or in other ways) if they smoke cannabis regularly? (Question C36c) | No risk (1)<br>slight risk (2)<br>moderate risk (3)<br>great risk (4) |

**Supplemental Table 3. Association between regular/high-risk cannabis use and all variables, bivariate analysis.**

| | Regular use | | | High-risk use | | |
|---|---|---|---|---|---|---|
| | % | OR | 95% CI | % | OR | 95% CI |
| **Sex** | | | | | | |
| Female | 16.2 | 1 | | 32.6 | 1 | |
| Male | 22.7 | 1.53 | (1.35-1.73) | 34.7 | 1.10 | (1.02-1.19) |
| **Father education** | | | | | | |
| Completed college or university | 17,1 | 1 | | 28.9 | 1 | |
| Completed primary school or less | 24,2 | 1.55 | (1.20-2.00) | 41.9 | 1.77 | (1.49-2.11) |
| Some secondary school | 21,2 | 1.30 | (1.05-1.62) | 36.0 | 1.38 | (1.20-1.58) |
| Completed secondary school | 18,6 | 1.11 | (0.93-1.32) | 33.1 | 1.22 | (1.09-1.36) |
| Some college or university | 17,9 | 1.06 | (0.85-1.31) | 33.3 | 1.23 | (1.07-1.4) |
| **Mother education** | | | | | | |
| Completed college or university | 16,7 | 1 | | 29.9 | 1 | |
| Completed primary school or less | 25,8 | 1.73 | (1.30-2.30) | 42.0 | 1.69 | (1.40-2.06) |
| Some secondary school | 21,7 | 1.38 | (1.10-1.73) | 38.5 | 1.47 | (1.27-1.69) |
| Completed secondary school | 19,2 | 1.18 | (1.00-1.40) | 33.8 | 1.20 | (1.08-1.33) |
| Some college or university | 19,7 | 1.22 | (1.00-1.48) | 32.9 | 1.15 | (1.01-1.30) |
| **Availability: Get cannabis** | | | | | | |
| Impossible or very difficult | 19,8 | 1 | | 26.0 | 1 | |
| Fairly difficult | 7,1 | 0.31 | (0.19-0.50) | 20.5 | 0.74 | (0.58-0.94) |
| Fairly easy | 11,0 | 0.50 | (0.36-0.69) | 27.1 | 1.06 | (0.87-1.30) |
| Very easy | 27,3 | 1.53 | (1.13-2.07) | 44.0 | 2.24 | (1.84-2.73) |
| **Risk: Try Cannabis** | | | | | | |
| Great risk | 24,8 | 1 | | 42.6 | 1 | |
| No risk | 22,1 | 0.86 | (0.63-1.16) | 35.2 | 0.73 | (0.60-0.90) |
| Slight risk | 13,8 | 0.48 | (0.35-0.67) | 29.4 | 0.56 | (0.45-0.69) |
| Moderate risk | 15,4 | 0.55 | (0.37-0.83) | 35.8 | 0.75 | (0.59-0.96) |
| **Risk: Cannabis occasionally** | | | | | | |
| Great risk | 17,0 | 1 | | 35.5 | 1 | |
| No risk | 26,7 | 1.78 | (1.33-2.38) | 41.5 | 1.29 | (1.09-1.53) |
| Slight risk | 16,1 | 0.94 | (0.69-1.26) | 30.8 | 0.81 | (0.68-0.96) |
| Moderate risk | 12,4 | 0.69 | (0.50-0.96) | 27.6 | 0.69 | (0.58-0.83) |
| **Risk: Cannabis regularly** | | | | | | |
| Great risk | 11,7 | 1 | | 27.2 | 1 | |
| No risk | 35,2 | 4.10 | (3.39-4.96) | 49.7 | 2.65 | (2.35-2.99) |
| Slight risk | 20,4 | 1.93 | (1.59-2.35) | 38.2 | 1.65 | (1.48-1.85) |
| Moderate risk | 15,3 | 1.36 | (1.12-1.66) | 29.9 | 1.14 | (1.03-1.27) |

**Supplemental Table 4. Association between perceived socioeconomic status and mediators (perceived availability/risk of use)**

| | Difficult: Get cannabis | | | | Risk: Try Cannabis | | | | Risk: Cannabis occasionally | | | | Risk: Cannabis regularly | | | |
|---|---|---|---|---|---|---|---|---|---|---|---|---|---|---|---|---|
| **Father education** | **Very difficult** | **Fairly difficult** | **Fairly easy** | **Very easy** | **No risk** | **Slight risk** | **Mod. risk** | **Great risk** | **No risk** | **Slight risk** | **Mod. risk** | **Great risk** | **No risk** | **Slight risk** | **Mod. risk** | **Great risk** |
| Completed primary school or less | 52.9 | 12.3 | 20.2 | 14.6 | 24.5 | 28.9 | 21.2 | 25.4 | 13.8 | 18.8 | 32.0 | 35.5 | 10.4 | 6.8 | 15.0 | 67.7 |
| Some secondary school | 44.9 | 14.4 | 25.5 | 15.3 | 23.2 | 30.9 | 21.6 | 24.3 | 11.5 | 23.1 | 33.6 | 31.8 | 6.1 | 7.6 | 19.6 | 66.8 |
| Completed secondary school | 47.3 | 14.7 | 23.9 | 14.0 | 20.8 | 28.9 | 21.5 | 28.8 | 10.1 | 20.4 | 33.1 | 36.4 | 5.1 | 7.0 | 17.9 | 69.9 |
| Some college or university | 45.8 | 14.6 | 24.2 | 15.5 | 21.5 | 30.5 | 22.2 | 25.8 | 9.8 | 22.1 | 34.8 | 33.3 | 5.1 | 7.4 | 20.1 | 67.5 |
| Completed college or university | 40.7 | 16.9 | 26.7 | 15.6 | 23.8 | 32.0 | 21.2 | 22.9 | 10.3 | 23.0 | 35.1 | 31.6 | 5.1 | 7.5 | 19.3 | 68.1 |
| Total | 45.1 | 15.2 | 24.8 | 14.9 | 22.5 | 30.4 | 21.5 | 25.6 | 10.6 | 21.7 | 34.0 | 33.7 | 5.6 | 7.3 | 18.7 | 68.4 |
| **Mother education** | **Very difficult** | **Fairly difficult** | **Fairly easy** | **Very easy** | **No risk** | **Slight risk** | **Mod. risk** | **Great risk** | **No risk** | **Slight risk** | **Mod. risk** | **Great risk** | **No risk** | **Slight risk** | **Mod. risk** | **Great risk** |
| Completed primary school or less | 57.6 | 11.5 | 18.3 | 12.7 | 23.5 | 26.3 | 20.9 | 29.2 | 13.5 | 16.9 | 31.0 | 38.6 | 10.7 | 5.3 | 14.2 | 69.8 |
| Some secondary school | 47.5 | 13.9 | 23.2 | 15.5 | 22.7 | 30.1 | 22.0 | 25.2 | 11.8 | 22.9 | 33.3 | 32.0 | 6.6 | 7.8 | 19.6 | 66.0 |
| Completed secondary school | 47.0 | 14.4 | 24.1 | 14.5 | 21.1 | 29.2 | 21.5 | 28.3 | 10.1 | 20.6 | 33.3 | 36.0 | 5.3 | 6.9 | 18.0 | 69.8 |
| Some college or university | 44.8 | 14.5 | 24.5 | 16.3 | 21.6 | 31.1 | 22.7 | 24.6 | 9.9 | 22.4 | 35.4 | 32.3 | 5.3 | 7.3 | 19.7 | 67.7 |
| Completed college or university | 41.2 | 16.9 | 26.9 | 15.0 | 23.8 | 32.1 | 21.4 | 22.8 | 10.5 | 23.0 | 34.9 | 31.5 | 5.0 | 7.8 | 19.3 | 67.9 |
| Total | 45.1 | 15.2 | 24.8 | 14.9 | 22.5 | 30.5 | 21.6 | 25.3 | 10.6 | 21.8 | 34.1 | 33.5 | 5.6 | 7.3 | 18.7 | 68.4 |

Legend: Percentages are calculated within each category of the SES variables (row percentages). The Pearson chi-square tests were used to compare categorical variables and all associations were statistically significant (p-value <0.0001). Mod.: moderate.

**Supplemental Figure 1. Mediation analysis of outcomes of cannabis use: current regular cannabis (A and B) use and past year high-risk use (C and D).**

**A**

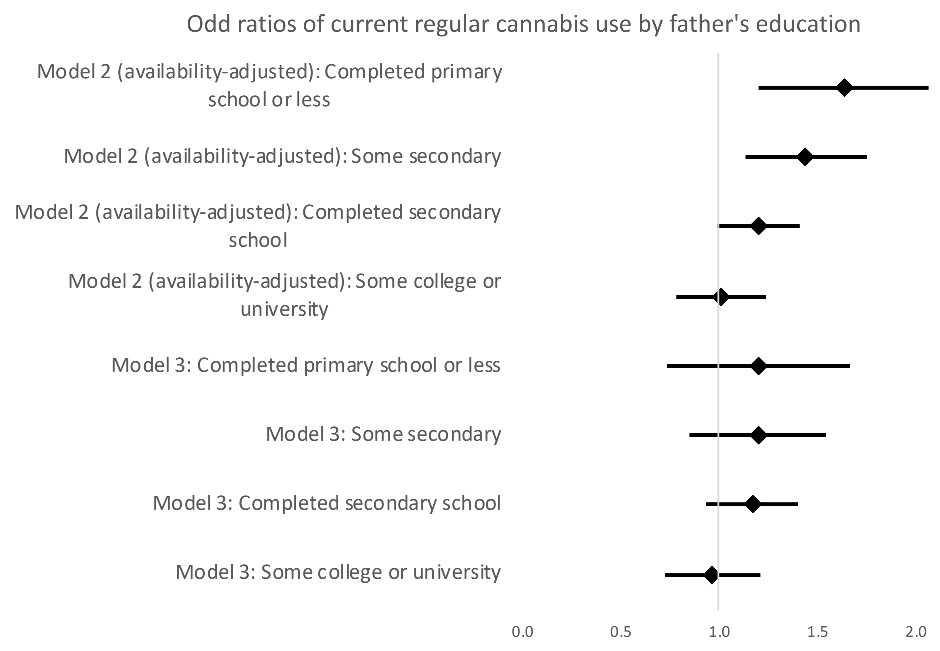


**B**

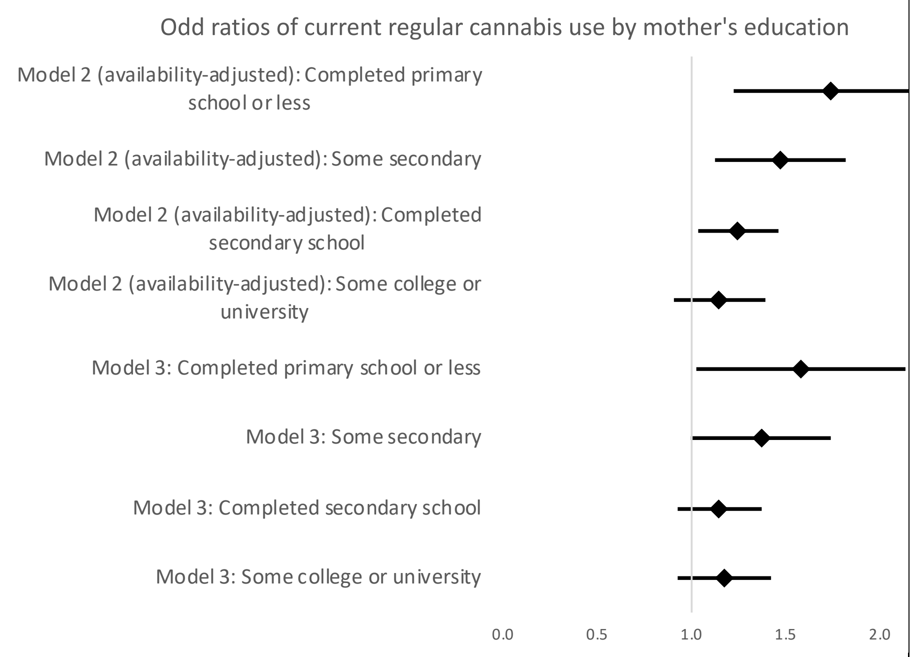


C

D

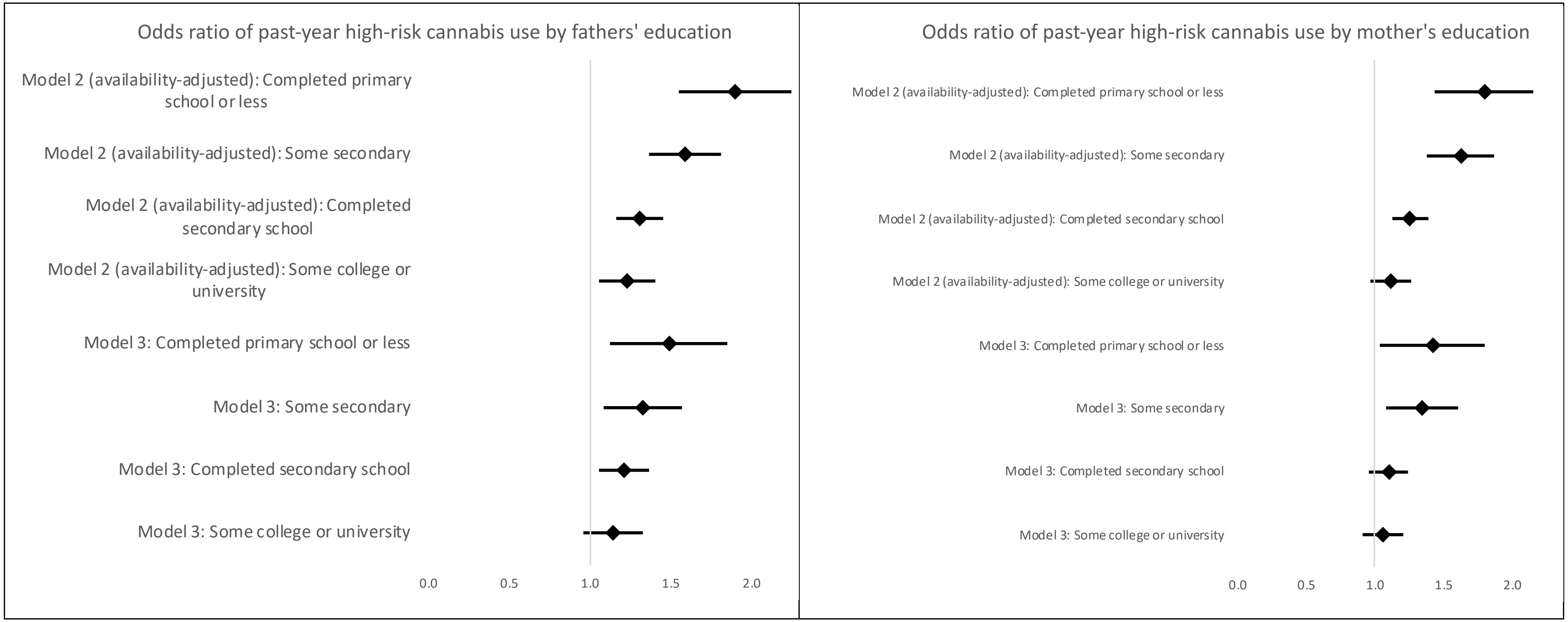


Legend: The highest level of education (completed college or university) was used as reference category. This figure represents the results of the mediation analysis showed in Table 3 and 4.
